# Diagnostic performance of serum CA-125 for overall and complicated acute appendicitis: a systematic review and meta-analysis

**DOI:** 10.1101/2023.12.16.23300068

**Authors:** Javier Arredondo Montero, Blanca Paola Pérez Riveros, Oscar Emilio Bueso Asfura

## Abstract

**Background:** This study aimed to analyze the diagnostic performance of serum CA-125 in acute appendicitis (AA).

**Methods:** This review was registered in PROSPERO (CRD42023450988). We included prospective or retrospective original clinical studies evaluating the diagnostic performance of serum CA-125 in AA. A search was conducted in PubMed, Web of Science, Scopus, and OVID. Search terms and keywords were: (appendicitis OR appendectomy) AND (CA-125 OR CA125). Two independent reviewers selected the articles and extracted relevant data. Methodological quality was assessed using the QUADAS2 index. A synthesis of the results, standardization of the metrics, and three random-effect meta-analyses were performed.

**Results:** Five studies with data from 533 participants (including 219 patients with a confirmed diagnosis of AA and 107 controls) were included in this review. The random-effect meta-analysis of serum CA-125 (AA vs controls) included 5 articles (125 AA and 70 controls) and resulted in a non-significant mean difference [95% CI] of −6.80 [−20.51,6.92] U/mL (p=0.33). The meta-analysis by subgroups that included only male patients resulted in a significant mean difference [95% CI] of 3.48 [0.46,6.49] U/mL (p=0.02)

**Conclusions:** Although serum CA-125 does not appear to be a good overall marker for the diagnosis of AA, our subgroup analyses show that this marker could be useful for diagnosing AA in males. It also appears to be a potentially useful tool for discriminating complicated and uncomplicated AA. However, the limited number of included studies precludes drawing generalizable conclusions. Future prospective studies focused on males are required to confirm these findings.

**Funding:** None

**Registration:** PROSPERO (CRD42023450988).

## Introduction

Acute appendicitis (AA) is the most prevalent urgent abdominal surgical pathology in the world [1]. AA remains a complex entity to diagnose, given the great variability of its clinical presentation, the broad differential diagnosis of acute abdomen, and the absence of perfect diagnostic tests [1,2]. Research developed in recent years regarding this matter has identified promising diagnostic biomarkers, such as interleukin 6 (IL-6) or serum amyloid A1 (SAA1) [3,4]. It should be noted, however, that these markers often have high processing times and economic costs and are not available in all Emergency Departments. In this sense, it is pertinent to highlight the role of the ratios derived from the complete blood count (CBC) such as platelet-to-lymphocyte ratio (PLR) or systemic immune inflammation index (SII), due to their ease of implementation, fast processing time and low economic cost [5,6]. Although these lines of research are of great interest, at present none of these biomarkers or ratios have reached a diagnostic performance that would allow them to be considered unique diagnostic tests.

CA-125 is a high molecular weight glycoprotein that can be produced by different tissues of the human body, particularly by the mesothelia (pleura, peritoneum, and pericardium) [7]. Although its most defined role in clinical practice is as an ovarian tumor marker, it is known that it is a marker that can be elevated in multiple pathophysiological contexts, especially in those that condition inflammation on the previously mentioned mesothelia. Under this premise and given that AA is invariably associated with inflammation of the peritoneum, we hypothesized that serum CA-125 could be a marker of interest for the diagnosis of AA. The objective of this systematic review is to characterize the diagnostic performance of serum CA-125 in AA.

## Methods

### Literature search and selection

We followed the Preferred Reporting Items for Systematic Reviews and Meta-Analyses in Diagnostic Test Accuracy Studies (PRISMA-DTA) guidance [8]. We specifically designed and implemented a review protocol that was registered in the international prospective register of systematic reviews (PROSPERO ID CRD42023450988). Eligible studies were identified by searching the main existing medical bibliography databases (PubMed, Web of Science, Scopus, OVID). Search terms and keywords were: (appendicitis OR appendectomy) AND (CA125 OR CA-125). The search was last executed on 15.10.2023.

Inclusion and exclusion criteria are shown in Supplementary File 1. JAM and BPR made the selection of articles by using the COVIDENCE ® tool. The results of the searches were imported into the platform and the articles were screened separately by both authors. Disagreements were resolved by consensus.

### Quality assessment

The selected articlés methodological quality and risk of bias were evaluated with the QUADAS2 tool [9]. Patient selection, index test, reference standard, and flow and timing were evaluated in each selected article. Applicability concerns regarding patient selection, index tests, and reference standards were also assessed.

### Data extraction and synthesis

Target condition was defined as histopathologically confirmed AA or formal clinical suspicion of AA. Index test was defined as serum CA-125. Reference standard was defined as the histopathological study of the cecal appendix. Three independent reviewers (JAM, BPR, OBA) extracted the relevant data from the selected articles following a standardized procedure. Extracted data included author, country where the study was conducted, year of publication, study design, study population (sample size, age range, and sex distribution), AA group and control group (CG) definitions, reference standard used in AA group, mean and standard deviation (or median and interquartile range) for serum CA-125, statistical p-value for the between-group comparison, serum CA-125 area under the curve (AUC), serum CA-125 cut-off value (if established), and its associated sensitivity and specificity. There were no disagreements between the reviewers after collating the extracted data. The metrics used in each study were reviewed and it was determined that a standardization of units was not required. Medians (Interquartile ranges) were converted to means (standard deviations) following a standardized procedure [10].

### Meta-analysis

Three random-effects meta-analyses (MA) for serum CA-125 (AA vs control group) were performed: 1) MA including all available studies. 2) MA including only studies with a male population 3) MA including only studies with a sex-stratified population (i.e. not mixed populations). A subgroup analysis for the studies including only a female population was not performed because there was only one study that met these characteristics. The results were expressed as a mean difference (95% CI) and were plotted in a forest plot. Between-study heterogeneity was assessed using the Chi^2^ and I^2^ statistics.

## Results

The search returned 138 articles (Scopus n=6; Pubmed n=53; Web of Science n=46; OVID n=33). Fifty-five duplicates were removed. Among the remaining 83 articles, we excluded 78 (inclusion and exclusion criteria, n=77; reports not retrieved, n=1). This review finally included 5 studies with data from 533 participants (including 219 patients with a confirmed diagnosis of AA and 107 controls) [11–15]. The lack of concordance between the sample size of the AA and control groups and the total sample size of the review is because not all studies clarified this information. The flowchart of the search and selection process is shown in Figure 1.

**Figure 1.**
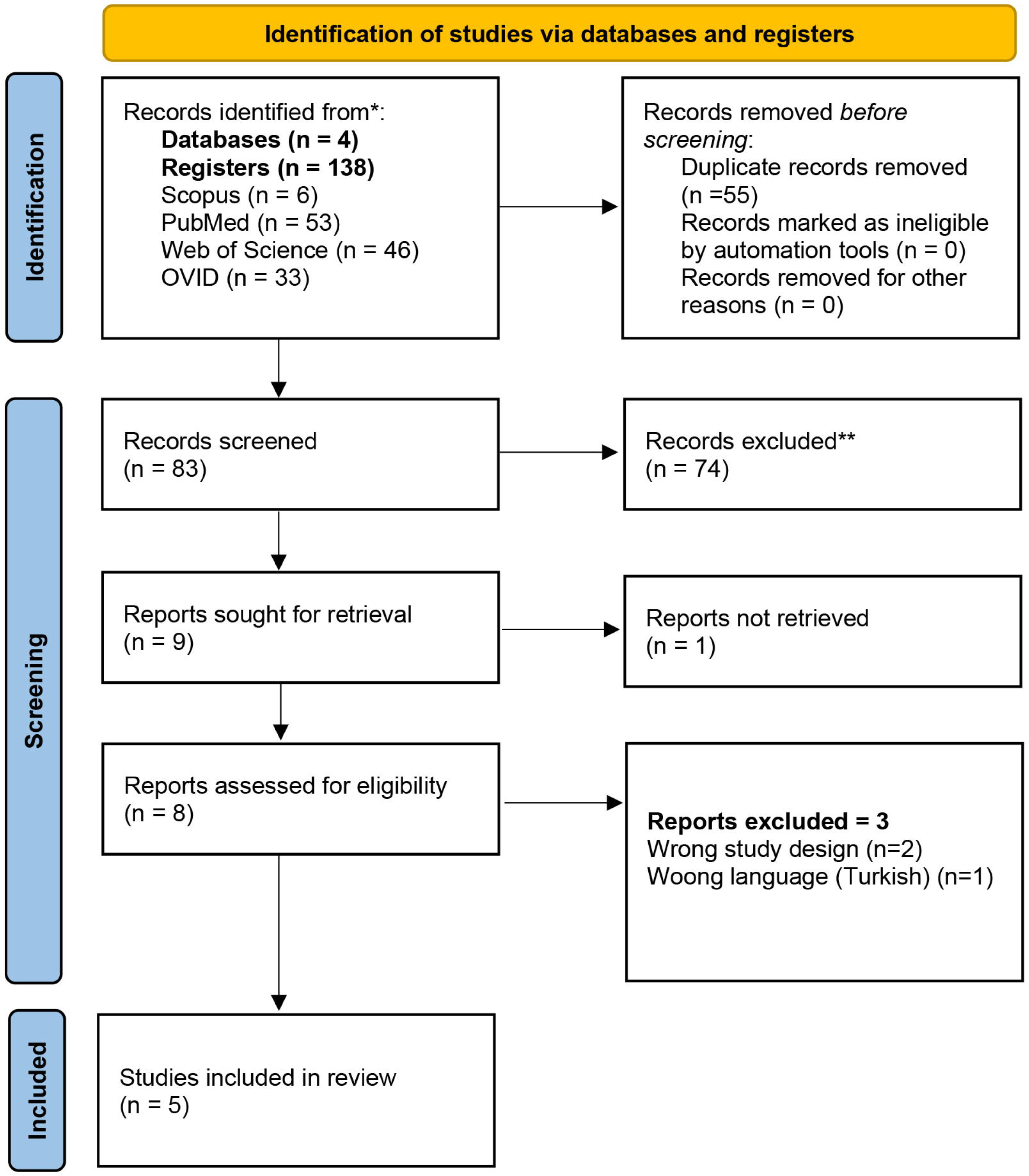
Flowchart of the search and selection process.

The risk of bias concerning the selection of patients was considered low in two of the studies [12,14] and unclear in three of them [11,13,15]. The risk of bias concerning the index test was considered low in four of the studies [11–14] and unclear in one of them [15]. The risk of bias concerning the reference standard was considered low in four of the studies [11–14] and unclear in one of them [15]. The risk of bias concerning flow and timing was considered low in two of the studies [11,14], unclear in two of the studies [12,13] and high in one of them [15]. Regarding applicability concerns, the risk was estimated as low in all articles except for Ahmadinejad et al., which was found to be unclear in all three categories [15]. The results of the QUADAS2 analysis are shown in Figure 2.

**Figure 2.**
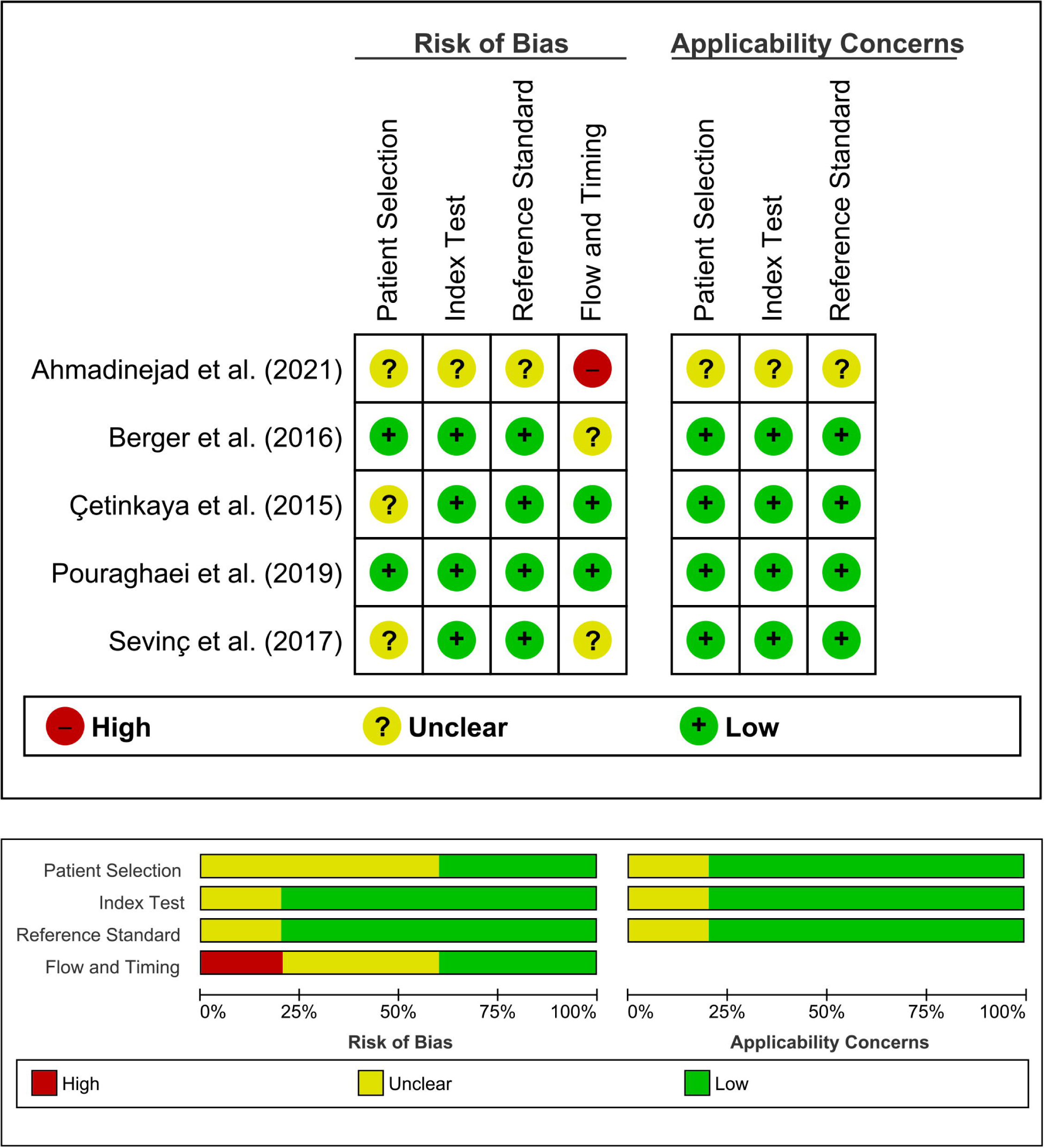
Graphical representation of the quality assessment of the diagnostic accuracy studies included in the review (QUADAS2).

### Serum CA-125 in acute appendicitis

#### Sociodemographic characteristics

The data extracted from the 5 studies that compared urinary serum CA-125 levels are summarized in Table 1. All studies were carried out between 2015 and 2021. One was from Israel [12], two from Turkey [11,13], and two from Iran [14,15]. All studies were prospective [11–15]. None of the studies involved pediatric populations. Three studies stratified their population and analyses by sex [12–14], and one of them included only a male population [13]. In all studies, cases were recruited in the Emergency Department before diagnosis, and biological samples were obtained at the time of inclusion in the study.

“The definition of “case” was consistent in four of the selected studies, given as the histopathological confirmation of AA in the surgical specimen [11–14]. One study did not provide sufficient data to determine this information [15]. This was not the case for the definition of “control”, which constituted either non-surgical abdominal pain [12], negative appendectomies [12,14], or healthy controls [13]. One study did not provide sufficient data to determine this information [15]. One study exclusively compared patients with uncomplicated AA (NCAA) and complicated AA (CAA) with no control group [11]. Three studies compared an AA group with a control group [12–14]. One study did not provide sufficient data to determine this information [15].

Three studies specifically evaluated potential conditions that could have altered serum CA-125 levels [12,14,15] including congestive heart failure, chronic lung disease, liver cirrhosis, inflammatory bowel disease, any malignancy, pregnancy (first trimester), endometriosis, and recent abdominal surgery. Sevinç et al. [13] opted to exclude the female population from the study, arguing that ovarian disorders and fluctuations during menstrual cycles could alter the results. Two studies [11,13] did not specify specific exclusions regarding possible confounders of CA-125 elevation.

#### CA-125 measurement units

Three studies reported the values as a mean (standard deviation) [12–14], 1 study as a median (interquartile range) [11], and one study did not provide quantitative values for 5-HIAA [15]. The reported measurement units for CA-125 were U/mL [11–15]. Following a standardized procedure, we converted the median (range) data Çetinkaya et al. [11] to mean (standard deviation) [10].

#### Determination of CA-125

Four studies determined CA-125 using electrochemiluminescence (ECL) immunoassay [11,12,14,15]. Specifically, one study used an UniCel DxI 800 system (Beckman Coulter, Brea, CA, USA) [12] and two used an E411 Cobas machine and kits produced by Germany Roche Company [14,15]. One study did not clarify the determination method [13].

### Serum CA-125 (AA vs CG)

Three studies compared an AA group with a control group [12–14]. <u>Diagnostic performance of serum CA-125 (AA vs CG)</u> Three studies provided a p-value for the comparison between CA-125 values in the AA group and in the control group, two of them being statistically significant [13,14]. The area under the curve (AUC) reported ranged from 0.62 [14] to 0.795 [13], and the proposed cut-offs ranged from 5.22 U/mL [13] to 16.4 U/mL [14,15]. The reported sensitivity ranged from 77.8% [14] to 81% [13] and the reported specificity ranged from 50% [14] to 65% [13].

#### Random-effects meta-analysis for serum CA-125

The random-effect meta-analysis for serum CA-125 (AA vs CG) included 5 articles (125 AA and 70 controls) and resulted in a non-significant mean difference [95% CI] of −6.80 [−20.51,6.92] U/mL (p=0.33). The I^2^ value was 96% and the Chi^2^ was 22.61. The subgroup meta-analysis that included only male patients resulted in a significant mean difference [95% CI] of 3.48 [0.46,6.49] U/mL (p=0.02). The I^2^ value was 61% and the Chi^2^ was 2.59. The subgroup meta-analysis that included only studies separated by sex (both male and female) resulted in a non-significant mean difference [95% CI] of 2.97 [−0.79,6.74] U/mL (p=0.12). The I^2^ value was 62% and the Chi^2^ was 5.22. The graphical depiction of the analyses is shown in Figure 3.

**Figure 3.**
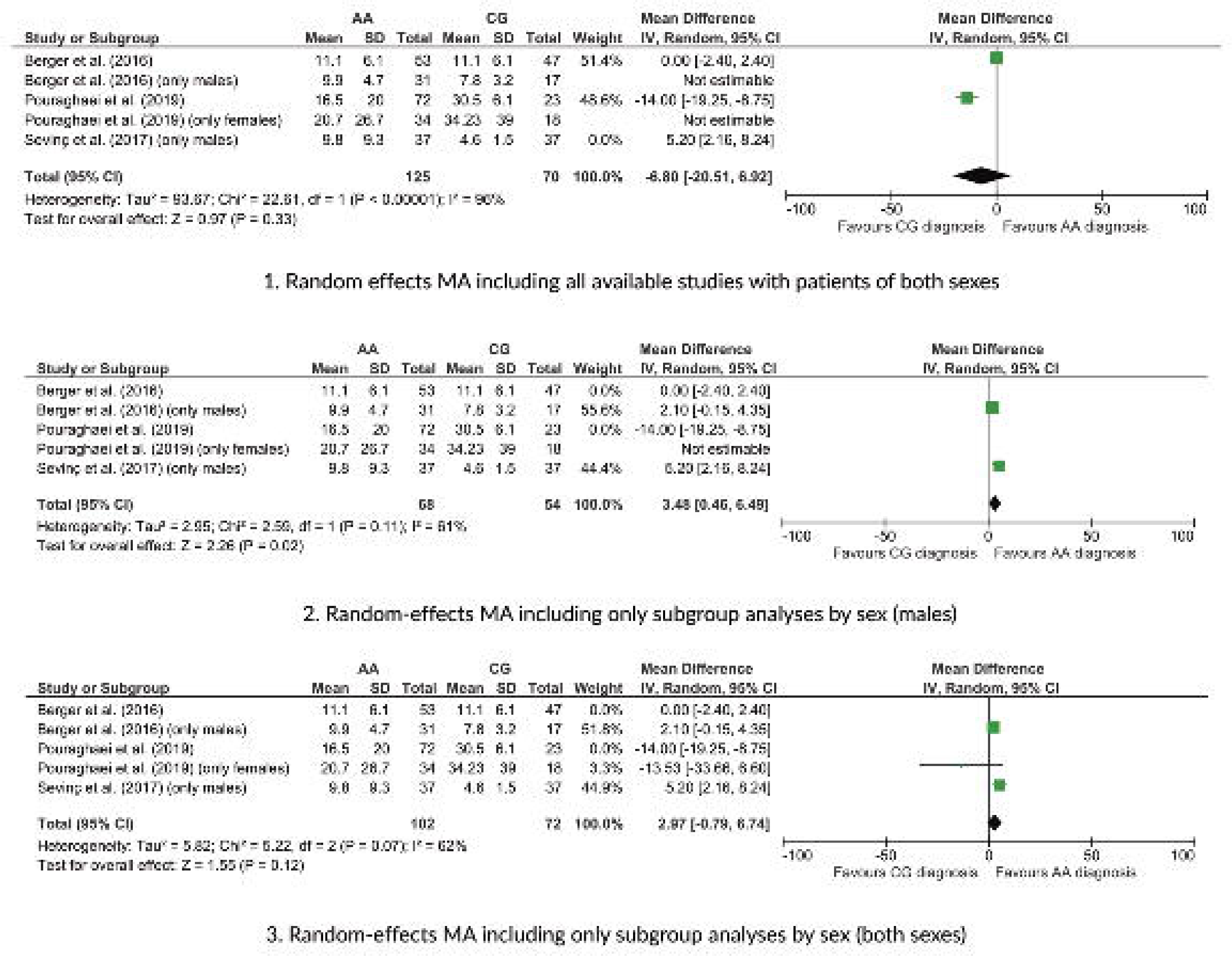
Forest plot of the random-effects meta-analyses performed for urinary 5-HIAA (AA group vs. Control group).

### Serum CA-125 (Complicated appendicitis vs non-complicated appendicitis)

Two studies compared the NCAA and CAA groups [11,12]. Çetinkaya et al. found a statistically significant difference between serum CA-125 levels in the NCAA and the CAA group [11], with an AUC of 0.885, a cut-off of 35, and a sensitivity and a specificity of 60% and 100% respectively. Berger et al. found a statistically significant correlation between the severity of AA in males and CA-125 levels [12] but they did not provide AUC values nor a cut-off and its sensitivity/specificity.

## Discussion

In 2010, A. Barasan published a paper postulating that CA-125 could be a useful marker for the diagnosis of AA [16]. He based his hypothesis on the in vitro peritonitis models developed by Zeillemaker et al. in 1994 [17], in which the authors documented that under various proinflammatory stimuli, the mesothelium produced CA-12 (the most effective stimulus being Interleukin-1). Zeillemaker et al. determined in this work that the time to secretion from the stimulus was approximately 6 hours. Wiwanitkit responded to this idea by raising the problem of standardization in the determination of this molecule (both in terms of laboratory methodology and the establishment of normality ranges for males and females) as well as the possible interference of the physiology and pathology of the female reproductive tract in this context.[18] Barasan subsequently argued that this molecule would indeed have a potentially higher diagnostic yield in males. During this exchange of correspondence, another interesting idea was raised, which was to perform serial determinations of this molecule (which could be an indirect measure of progressive peritoneal inflammation in acute appendicitis) [18]. This correspondence laid the foundation for all prospective studies included in this review.

In the present review, we analyzed the existing evidence regarding the role of serum CA-125 in the diagnosis of AA and the discrimination of NCAA and CAA. We synthesized the results of 5 prospective studies including 533 participants (219 patients with a confirmed diagnosis of AA and 107 controls). We performed three different meta-analyses regarding serum CA-125 values (AA vs CG): First, a random-effects meta-analysis including all available studies, which resulted in a non-significant mean difference between groups; second, a subgroup analysis for the studies conducted exclusively in male population, which did show a statistically significant mean difference between groups; and finally, a subgroup analysis for studies conducted in all populations stratified by sex (both male and female), which again showed a non-significant mean difference between groups. The CA-125 values obtained in the only-females study were higher in the CG group than in the AA group, contrary to what happened in the only-males studies. These findings, although based on a limited number of studies with small sample sizes, suggest that serum CA-125 could be a promising marker for the diagnosis of AA in males but that is not useful for the diagnosis of AA in the general population or females. One aspect that reinforces this analysis is the substantial decrease in heterogeneity in the analyses stratified by sex compared with the overall analysis (I^2^: 61% vs 96%).

Regarding the pathophysiological and etiopathogenic substrate underlying these findings, we believe it is consistent. The potential fluctuation in serum CA-125 values in reproductive-age female patients concerning menstrual cycles and the potential presence of concomitant gynecologic pathology support this premise. We believe it is pertinent to point out that of the 5 studies included, two did not adequately define solid exclusion criteria. Given the large number of situations in which there may be an elevation of CA-125, we believe it is imperative that future studies aimed at assessing this marker in diagnostic terms strictly apply such exclusion criteria.

Regarding the global diagnostic performance of serum CA-125 for the discrimination between AA and controls, there is a limited amount of data, although in general, the reported diagnostic yields are only moderate. The only study that reported diagnostic yields for the comparison between NCAA and CAA showed very promising values, with an AUC of 0.885. This coupled with the fact that the two studies that have evaluated the role of CA-125 in distinguishing NCAA from CAA showed statistically significant differences lead us to consider this line of research as promising for the future.

When we explored other potential sources of heterogeneity in this work, we found a relative homogeneity in molecule determination methodology. Except for one study that did not report the determination methodology, all determinations were performed through ECL. We believe that this is because CA-125 is an established diagnostic and prognostic molecule in healthcare practice. Regarding the study design, all the studies were prospective. We did find significant variability in the definition of the control group, which may constitute a source of heterogeneity. The use of a control group of healthy volunteers compared to a control group of non-surgical abdominal pain may overestimate the diagnostic performance of the molecule.

All the studies were conducted in Asian countries. it should be considered that the fact that all the studies were carried out in an Asian population prevents extrapolation of the findings to other populations without prior validation of the findings.

The main strengths of this work are its rigorous methodology, following international PRISMA-DTA guidelines, and the application of the QUADAS2 bias assessment tool [8,9]. Nevertheless, this review also has important limitations: 1) the number of studies included is very limited and the sample size included is small 2) not all the working groups have considered the potential factors that could have elevated serum CA-125 3) Due to the limited information reported by the different authors, it was not possible to perform a specific diagnostic performance meta-analytical model (such as DTA meta-analysis). 4) The finding of a significant mean difference between groups in a meta-analytic random-effects model does not invariably translate into adequate diagnostic performance, so the findings reported here should be interpreted with caution.

In conclusion and although the evidence is scarce, CA-125 should be considered a potential diagnostic biomarker in AA in males. Likewise, CA-125 could be a promising tool to discriminate between NCAA and CAA. In this last scenario, it remains to be elucidated whether the marker could be useful in the female population or not. There is no evidence of the diagnostic role of this molecule in the pediatric population. No one has evaluated, to our knowledge, the role of serial CA-125 determinations proposed by Barasan. We believe this is also an interesting line for the future. Future prospective studies with a large sample size. with adequate gender stratification and an adequate assessment of factors that may elevate CA-125 levels should be conducted.

## Supporting information

PRISMA-DTA checklist

## Data Availability

All data produced in the present study is available through the corresponding author.

