## Supplementary File 1 for "Diagnostic performance of serum CA-125 for overall and complicated acute appendicitis: a systematic review and meta-analysis"

**Exclusion criteria**

-Case reports.

-Duplicate or overlapping studies.

-Reviews, systematic reviews, consensus guidelines.

-Languages other than English or Spanish.

-Studies with no surgical intervention.

-Studies with no population of interest.

-Studies conducted in immunocompromised patients.

-Studies conducted in patients with metastatic neoplastic disease and invasive abdominal or gynecological neoplastic disease.

-Studies conducted in patients with acute or chronic kidney disease.

-Studies conducted in patients with gynecological pathology (endometriosis, pelvic inflammatory disease, non-cancerous ovarian cysts, uterine fibroids…).

Studies conducted in pregnant patients-

-Studies which analyzed CA-125 in any other biological sample than serum-

S

**Inclusion criteria**

-Prospective or retrospective observational original clinical studies evaluating the diagnostic accuracy of serum CA-125 in relation to the reference standards for the diagnosis of appendicitis and/or for the discrimination between complicated and uncomplicated appendicitis.

-Diagnostic validation studies evaluating the diagnostic accuracy of serum CA-125 in relation to the reference standards for the diagnosis of appendicitis and/or for the discrimination between complicated and uncomplicated appendicitis.

**Supplementary file 1. Inclusion and exclusion criteria**
