## Supplementary material for "Diagnostic performance of serum CA-125 for overall and complicated acute appendicitis: a systematic review and meta-analysis": Table 1.

| **Author** | **Country** | **Study design** | **Age (range)** | **Sex M/F** | **Total*N*** | ***N* in AA** | ***N* in CG** | **Serum CA-125 AA**  **(U/mL)** | **Serum CA-125 CG (U/mL)** | **P**  **(CG vs AA)** | **P (NCAA vs CAA)** | **Cut-off U/mL (CG vs AA)** | **AUC (AA vs control)** | **Sensitivity (%)** | **Specificity (%)** |
| --- | --- | --- | --- | --- | --- | --- | --- | --- | --- | --- | --- | --- | --- | --- | --- |
| Çetinkaya et al. (2015) | Turkey | Prospective | 18-60 | 35/22 | 57 | 57  **CAA:** 10 | - | **NCAA**: 10.5 (7.4-13.2)^2^  **NCAA:** 10.4 (4.4)^3^  **CAA:** 49.9 (14.1-71.2)^2^  **CAA:** 45.1 (49.1)^3^ | - | 0.001 | 0.001 | **NCAA vs CAA:** 35 | **NCAA vs CAA:** 0.885 | 60% | 100% |
| Berger et al. (2016) | Israel | Prospective | 18-50 | 48/52 | 100 | 53 | 47 | 11.1 (6.1)^1^  Only males: 9.9 (4.7)^1^ | 11.1 (6.1)^1^  Only males: 7.8 (3.2)^1^ | 0.97  Only males: 0.098 | 0.008*  0.02** | 8.75 | - | - | - |
| Sevinç et al. (2017) | Turkey | Prospective | 27.7 (8.4)^1^ | 74/0 (Only males included) | 74 | 37  **CAA**: 15 (10 **GA**, 5 **PA**) | 37 | Only males: 9.8 (9.3)^1^ | Only males: 4.6 (1.5)^1^ | 0.001 | - | 5.22 | 0.795 | 81% | 65% |
| Pouraghaei et al. (2019) | Iran | Prospective | 15-70 | 43/52 | 95 | 72  **CAA:** 23 | 23 | 16.5 (20.0)^1^  Only females: 20.7 (26.7)^1^ | 30.5 (6.1)^1^  Only females: 34.23 (39)^1^ | 0.001 | 0.058 | 16.4 | 0.62 | 77.8% | 50% |
| Ahmadinejad et al. (2021) | Iran | Prospective | 26.5 (1)^1^ | 101/206 | 207 | - | - | - | - |  | - | 16.4 | - | - | - |

**Table 1. Summary of publications included in this review.**

**AA:** Acute appendicitis group, **CG**: Control group, **NCAA**: Non-complicated acute appendicitis, **CAA:** Complicated acute appendicitis, **NS**: non-statistically significant **GA**: Gangrenous appendicitis; **PA**: perforated appendicitis

**1**: Mean (standard deviation) **2:** Median (Interquartile range) **3:** Mean (standard deviation) calculated from Median (Interquartile range)

**: Regression model establishing the correlation between an elevated preoperative CA-125 level in males and the likelihood of encountering severe appendicitis during surgery* ***: Regression model establishing the correlation between an elevated preoperative CA-125 level in males and the likelihood of diagnosing severe appendicitis during the histopathology study*
